# Impact of COVID-19 on mortality in coastal Kenya: a longitudinal open cohort study

**DOI:** 10.1101/2022.10.12.22281019

**Authors:** M Otiende, A Nyaguara, C Bottomley, D Walumbe, G Mochamah, D Amadi, C Nyundo, EW Kagucia, AO Etyang, IMO Adetifa, SPC Brand, E Maitha, E Chondo, E Nzomo, R Aman, M Mwangangi, P Amoth, K Kasera, W Ng’ang’a, E Barasa, B Tsofa, J Mwangangi, P Bejon, A Agweyu, TN Williams, JAG Scott

## Abstract

**Background:** There is uncertainty about the mortality impact of the COVID-19 pandemic in Africa because of poor ascertainment of cases and limited national civil vital registration. We analysed excess mortality from 1^st^ January 2020-5^th^ May 2022 in a Health and Demographic Surveillance Study in Coastal Kenya where the SARS-CoV-2 seroprevalence reached 75% among adults in March 2022 despite vaccine uptake of only 17%.

**Methods:** We modelled expected mortality in 2020-2022 among a population of 306,000 from baseline surveillance data between 2010-2019. We calculated excess mortality as the ratio of observed/expected deaths in 5 age strata for each month and for each national wave of the pandemic. We estimated cumulative mortality risks as the total number of excess deaths in the pandemic per 100,000 population. We investigated observed deaths using verbal autopsy.

**Findings:** We observed 16,236 deaths among 3,410,800 person years between 1^st^ January 2010 and 5^th^ May 2022. Across 5 waves of COVID-19 cases during 1st April 2020-16^th^ April 2022, population excess mortality was 4.1% (95% PI -0.2%, 7.9%). Mortality was elevated among those aged ≥65 years at 14.3% (95% PI 7.4%, 21.6%); excess deaths coincided with wave 2 (wild-type), wave 4 (Delta) and wave 5 (Omicron BA1). Among children aged 1-14 years there was negative excess mortality of -20.3% (95% PI -29.8%, -8.1%). Verbal autopsy data showed a transient reduction in deaths from acute respiratory infections in 2020 at all ages. For comparison with other studies, cumulative excess mortality risk for January 2020-December 2021, age-standardized to the Kenyan population, was 47.5/100,000.

**Interpretation:** Net excess mortality during the pandemic was substantially lower in Coastal Kenya than in many high income countries. However, adults, aged ≥65 years, experienced substantial excess mortality suggesting that targeted COVID-19 vaccination of older persons may limit further COVID-19 deaths by protecting the residual pool of naive individuals.

**Funding:** Wellcome Trust

## INTRODUCTION

Estimates of global excess mortality during the COVID-19 pandemic in 2020-2021 vary widely from 14.9 million to 19.7 million^1-3^. An important factor driving this variation is uncertainty regarding the impact of COVID-19 in Africa. Across sub-Saharan Africa, only South Africa contributed national mortality data to these models and the estimates of deaths in all other countries were extrapolated from mortality patterns observed elsewhere. This lack of data has led to controversy in the interpretation of the pandemic’s impact in Africa and this has significant consequences for policy. For example, predictions of a lower mortality impact in Africa, based on its youthful population structure, stimulated arguments to sustain health spending on existing threats such as malaria, HIV and respiratory tract infections in children rather than redirect funding to COVID-19 response measures^4^.

Because most African countries did not have comprehensive national civil vital registration systems at the time of the pandemic, this controversy cannot be fully resolved. Across the continent, however, there are numerous health and demographic surveillance systems (HDSS) which undertake longitudinal population-based mortality surveillance^5^. These cover only a fraction of the national population and cannot be considered representative of the whole country, but they provide a robust empiric insight into the longitudinal mortality experience of selected African populations throughout the pandemic.

The global number of COVID-19-confirmed deaths reached 5.4 million by December 2021 which is substantially smaller than the number modelled as excess mortality. Ascertainment of COVID-19 deaths was constrained by limited access to COVID-19 testing. Excess mortality, which sums deaths attributable both to COVID-19 and to the restrictions of the pandemic response, such as reduced access to health care, and which subtracts mortality gains attributable to the pandemic response, such as the suppression of influenza transmission, is considered a more useful measure of total impact^6-9^. It is calculated as a ratio: the total number of deaths observed in a population divided by the number of deaths expected on the basis of the population mortality experience in previous years. It can also be presented as a cumulative risk: the number of deaths in excess of expectation since the start of the pandemic divided by the size of the population under surveillance.

In the analyses presented here we estimate both measures of excess mortality in a HDSS population of 306,000 individuals in Kilifi, Kenya, which has been under continuous surveillance for over 20 years. The same HDSS was also used as a sampling frame for population-based studies of anti-SARS-CoV-2 antibodies. Seroprevalence among adults was 25% in December 2020 to April 2021^10^ and 75% in February to May 2022^11^ results in children were lower at 15% and 64%, respectively. Given only 17% of the adult population had been vaccinated in the second survey, this suggests wide dissemination of SARS-CoV-2 in Kilifi, typical of elsewhere in Africa^12-19^. Because excess mortality is dependent on the population age-structure we have reported age-stratified data^20^. We have estimated monthly excess mortality from 1^st^ April 2020 to 5^th^ May 2022, which includes the first five waves of SARS-CoV-2 in Kenya.

## METHODS

### Data source and study setting

The first case of COVID-19 in Kenya was identified on 12^th^ March 2020 and 323,818 cases and 5,649 deaths were reported up 5^th^ May 2022^21^. To date there have been six waves of COVID-19 in Kenya; the first two waves were driven by the wild-type variant, the third by the Beta/Alpha variants, the fourth by Delta and the fifth by Omicron BA1 (Figure 1).

**Figure 1.**
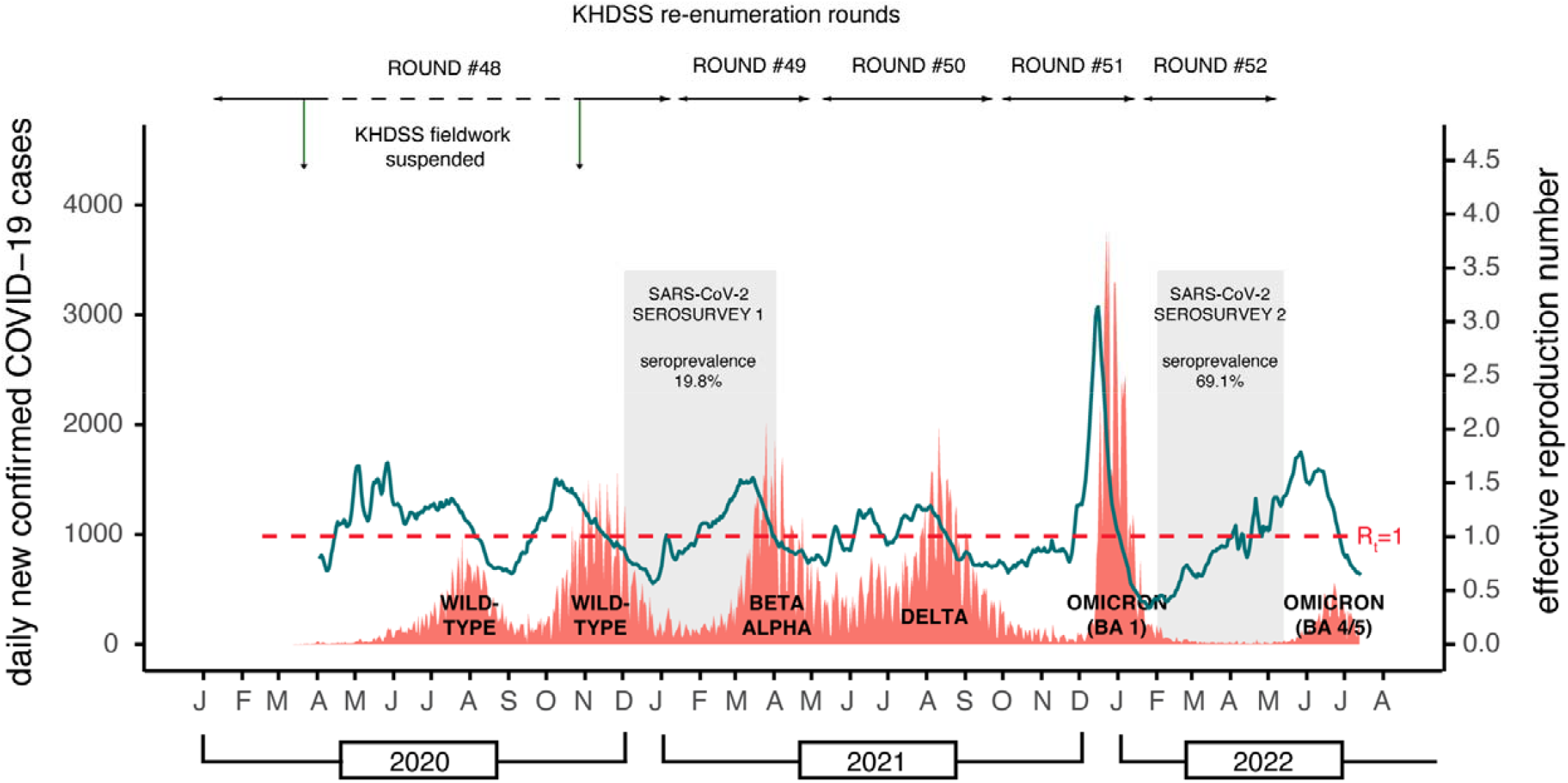
Timeline of the first six COVID-19 waves in Kenya, the Kilifi HDSS re-enumeration rounds and excess mortality analysis windows. Exact dates of the re-enumeration rounds, defined waves and analysis windows are listed in Table S5. The dotted horizontal line shows the period when HDSS fieldwork was suspended during round 48. The orange data series are the daily number of new cases of test-positive COVID-19 cases reported in Kenya (scale on left-hand y-axis) [Data source: https://coronavirus.jhu.edu/map.html]. The predominant variant behind each wave is denoted at the base of each wave. The green line represents the effective reproductive number (scale on right-hand y-axis). The dates of the two anti-SARS-CoV-2 antibody serosurveys in Kilifi HDSS are shown as grey bars^10,11^.

Kilifi HDSS was established in 2000^22^ with an initial census of 180,000 residents. Vital status and migration events have been recorded in subsequent re-enumeration rounds conducted every 4 months. By 2020, the population size was 306,000 and approximately 1300 deaths were recorded in each of the three years 2017-19^23^. In Kilifi, the first case of COVID-19 was detected on 22^nd^ March 2020. Because of a nationwide lockdown, HDSS field operations were suspended for 7 months between 23^rd^ March 2020 and 25^th^ October 2020.

Cause of death within Kilifi HDSS has been evaluated by Verbal Autopsy (VA) since 2008^24^. Trained field interviewers have questioned close relatives about signs and symptoms of the deceased household member from 30 days after the date of death. The interviews use the 2014 WHO VA questionnaire and are coded by the InterVA-4 algorithm^25^. From April 2020 we asked 6 additional questions (Table S1) that were based on WHO recommendations, intending to identify possible COVID-19 deaths^25^.

### Statistical analysis

#### Excess mortality

Mortality rates were calculated as the number of deaths divided by person years of observation (PYO). PYO were calculated as time from the latest of birth or in-migration or study start date to the earliest of death or outmigration or study end date. Individuals’ periods of residence outside the Kilifi HDSS area were excluded. Models fitted to monthly mortality counts from Jan 2010 to Dec 2019 were used to predict a counterfactual scenario for mortality in the absence of COVID-19 from January 2020 to April 2022. We chose January 2010 to December 2019 as the baseline period because mortality rates were stable during this decade^23^.

We modelled monthly death counts using negative binomial regression and computed the 95% prediction intervals from the 5^th^ and 95^th^ percentiles of 100 model simulations. The model included a log-linear trend, sine and cosine terms to account for seasonality, and an offset to account for changes in person years of observation (Equation S1). It was fitted using the glm.nb function in R.

Excess mortality was calculated as the difference between the observed and predicted number of deaths in the COVID-19 period expressed as a percentage of predicted number of deaths or as a cumulative risk based on the initial population size (per capita excess mortality). We used 1st April 2020 as the start of the analysis period during the pandemic because the interval from infection to death is approximately 2-3 weeks^26^ and it is unlikely there would have been any COVID-19 deaths before April 2020. We analysed overall and sex-specific excess mortality from 1^st^ January 2020 - 31^st^ March 2020, in each of the first 5 waves and for the entire duration. We defined the start of a wave as the point at which the daily effective reproduction^21,27^ number of SARS-CoV-2 traversed 1 in a positive direction after at least 4 weeks below 1 and the end of the wave to be the point where the subsequent wave begins. For excess mortality, analysis periods associated with each wave were offset by 2 weeks.

We locked our demographic database on 9^th^ September 2022, the date of completion of the 53^rd^ round and censored our analytic database on 5^th^ May 2022, the date our 52^nd^ HDSS re-enumeration round was completed. In the 10 re-enumeration rounds preceding the pandemic, 99% of deaths were ascertained by the end of the first complete re-enumeration round after the date of the death (Table S2). The remaining 1% were not detected until a second interview was undertaken because the initial respondent, who may have been a member of an adjacent household, may not have been aware of the death. To adjust for this under-ascertainment we inflated the number of deaths reported to have occurred during the 53^rd^ round (15^th^ Jan to 5^th^ May 2022) by 1%.

The Kilifi HDSS population is re-enumerated every 4 months because infant deaths may be missed with longer intervals; a child may be born and die without any enumeration contact. The suspension of the HDSS field operations on 23^rd^ March 2020 extended this re-enumeration interval from 4 to 11 months and it is likely that the ascertainment of infant deaths was reduced in this period. Therefore, we excluded infants from the estimation of overall excess mortality. Immigrants may also enter and die before they can be enumerated. To assess mortality changes attributable to variable detection of immigrant deaths, we compared the mortality risk of the snapshot cohort of Kilifi HDSS residents on 23^rd^ March in 2020 up to the 25^th^ October 2020 (the period that fieldwork was suspended) to similar snapshot cohort analyses beginning 23^rd^ March and terminating on 25^th^ October in each year of the baseline period, and in 2021.

Although fieldwork was suspended in 2020, deaths occurring in the suspension period were ascertained as soon as field studies resumed. However, even after the resumption of field activities, social restrictions may have limited access to some houses, thereby reducing the quality of information gathered; we examined this possibility by analysing the source of each re-enumeration record (household members vs neighbours) during the pandemic years compared to the baseline period.

#### Cause of death (verbal autopsy)

We calculated cause-specific mortality fractions in 10 categories; the 9 leading causes of death in children and adults and ‘all other causes’; the specific causes examined were Acute Respiratory Infections (ARI), unspecified cardiac disease, stroke, pulmonary tuberculosis, malaria, HIV-related deaths, digestive neoplasm, acute abdominal conditions, and Road Traffic Accidents (RTA).

For any death with at least one positive response to the 6 VA COVID-19 questions proposed by the WHO, we applied two discriminating processes. Firstly, we invited two independent reviewers to conduct Physician Certified Verbal Autopsy (PCVA) using clinical information collected during VA interview and classified each death as a probable, possible or unlikely COVID-19 death. Discordant cases were resolved jointly by the reviewers. Secondly, we used the COVID-19 Rapid Mortality Surveillance (CRMS) software^28^, which is a simplified version of the probabilistic modelling methods used in the InterVA-4 models, to derive the probability that the death was COVID-19 related. We used a probability cut-off value of 0.89 based on a validation study conducted in Brazil^29^.

## RESULTS

### Excess mortality

We analysed 16,236 deaths occurring between 1^st^ Jan 2010 and 5^th^ May 2022 from among 3,410,800 PYO. Observed and expected monthly mortality rates are shown in Figure S1 and excess mortality, derived from these data are shown in Figure 2. On aggregating all ages except infants, there was significant excess mortality in December 2020, July and August 2021, and December 2021. In age-specific analyses, there was significant excess mortality in December 2020 among those aged 15-45 years and ≥65 years; in July/August 2021 among those aged 45-65 years and ≥65 years; and in December 2021 among those aged ≥65 years. These periods coincide with the tail of wave 2 (wild-type), the rise and peak of wave 4 (Delta) and the peak of wave 5 (Omicron BA1). As might be anticipated by methodological constraints that limit the detection of deaths shortly after birth, observed excess mortality among infants was generally negative during the first year of the pandemic (Figure 2).

**Figure 2.**
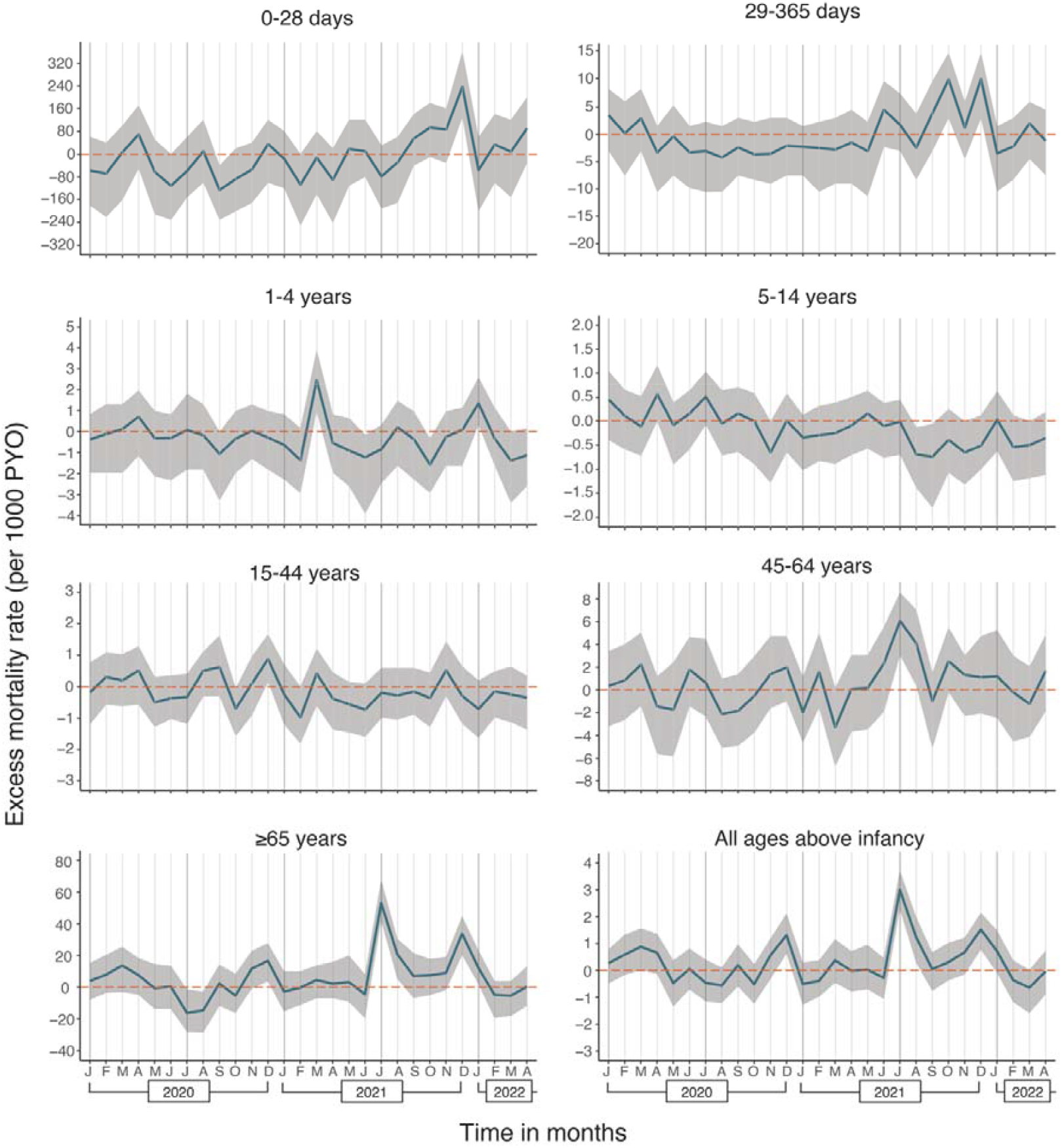
Monthly excess mortality rates from 1^st^ April 2020 – 30^th^ April 2022. Calculated as (observed deaths - expected deaths)/PYO. The excess mortality rate for all ages above infancy (age ≥1 year) is the weighted average of the age-specific rates. The weights are the proportion of each age-group in the Kilifi HDSS population.

In analyses by wave period, there was significant excess mortality for all ages together (excluding infants) during only one wave period, wave 4 (Delta, Table 1a). We predicted 609 deaths during the Delta wave but observed 711 deaths (excess mortality 16.7%, 95% PI 9.5%, 25.6%). In age-specific analyses, there was significant excess mortality in wave 2 (wild type) among those aged ≥65 years; in wave 4 (Delta) among those aged 45-64 years and ≥65 years; and in wave 5 (Omicron BA1) among those aged ≥65 years. Mortality was significantly lower than expected in wave 4 (Delta) among those aged 1-14 years; and in wave 5 (Omicron BA1) among those aged 5-14 years. Surprisingly, in the three months before the start of the pandemic (1^st^ January - 31^st^ March, 2020), we observed an overall excess mortality of 14.7% (95% PI 3.3%, 27.2%) (Table 1a). In age-specific analyses, this was significant only among those aged ≥65 years (22.6%, 95% PI 5.2%, 51.0%).

**Table 1a.**
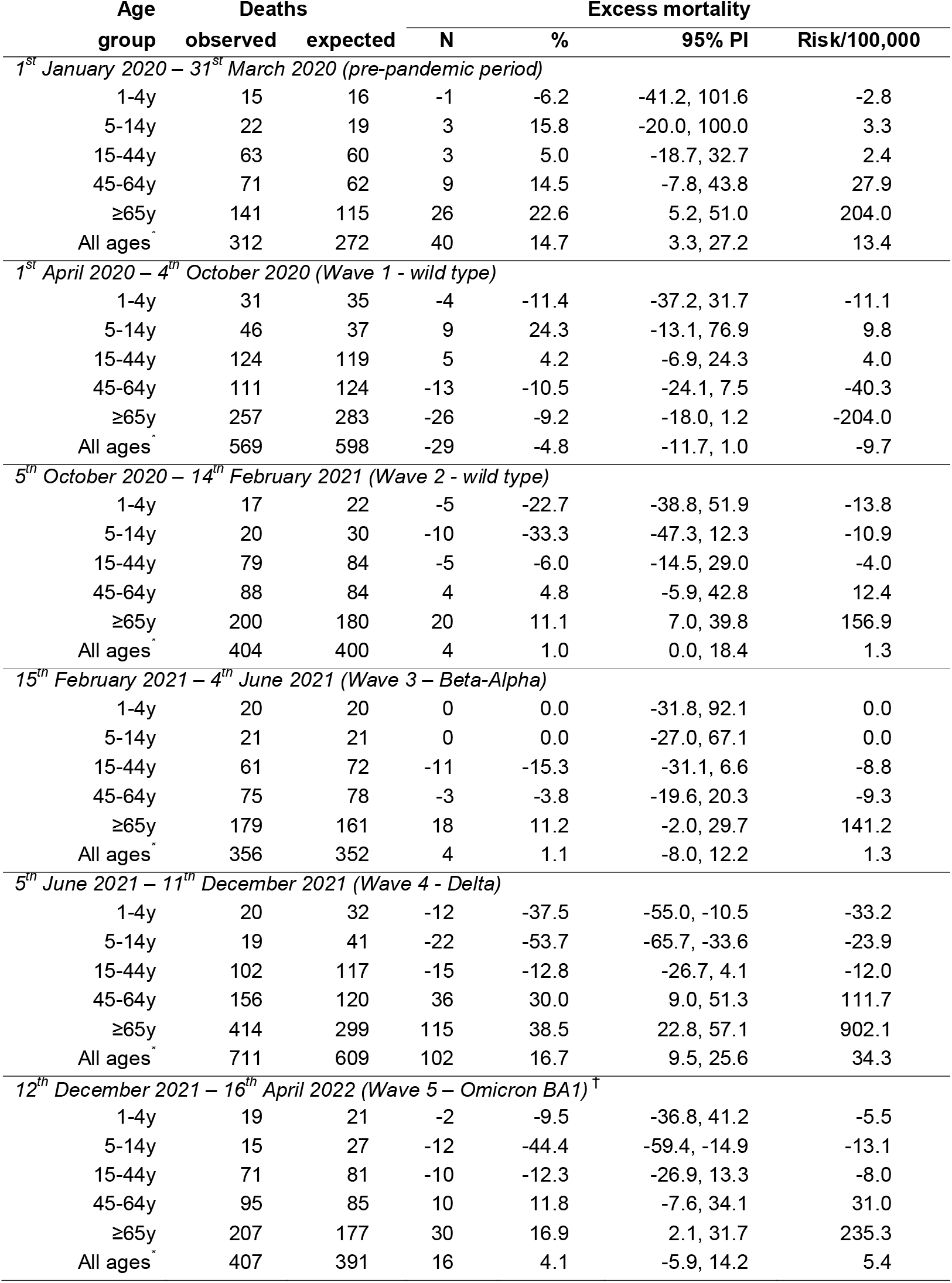
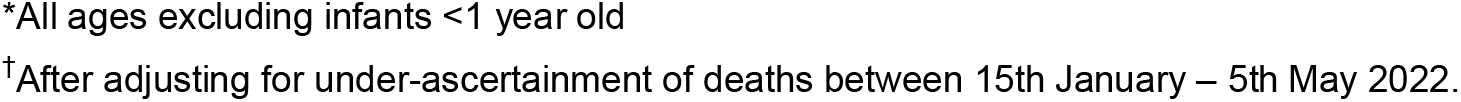
Excess deaths from 1^st^ January 2020 to 16^th^ April 2022 in Kilifi HDSS residents aged ≥1y, pre-pandemic and in each of 5 COVID-19 waves.

| Age group | Deaths |  | Excess mortality |  |  |  |
| --- | --- | --- | --- | --- | --- | --- |
|  | observed | expected | N | % | 95% PI | Risk/100,000 |
| *1<sup>st</sup> January 2020 – 31<sup>st</sup> March 2020 (pre-pandemic period)* |  |  |  |  |  |  |
| 1-4y | 15 | 16 | -1 | -6.2 | -41.2, 101.6 | -2.8 |
| 5-14y | 22 | 19 | 3 | 15.8 | -20.0, 100.0 | 3.3 |
| 15-44y | 63 | 60 | 3 | 5.0 | -18.7, 32.7 | 2.4 |
| 45-64y | 71 | 62 | 9 | 14.5 | -7.8, 43.8 | 27.9 |
| ≥65y | 141 | 115 | 26 | 22.6 | 5.2, 51.0 | 204.0 |
| All ages<sup>*</sup> | 312 | 272 | 40 | 14.7 | 3.3, 27.2 | 13.4 |
| *1<sup>st</sup> April 2020 – 4<sup>th</sup> October 2020 (Wave 1 - wild type)* |  |  |  |  |  |  |
| 1-4y | 31 | 35 | -4 | -11.4 | -37.2, 31.7 | -11.1 |
| 5-14y | 46 | 37 | 9 | 24.3 | -13.1, 76.9 | 9.8 |
| 15-44y | 124 | 119 | 5 | 4.2 | -6.9, 24.3 | 4.0 |
| 45-64y | 111 | 124 | -13 | -10.5 | -24.1, 7.5 | -40.3 |
| ≥65y | 257 | 283 | -26 | -9.2 | -18.0, 1.2 | -204.0 |
| All ages<sup>*</sup> | 569 | 598 | -29 | -4.8 | -11.7, 1.0 | -9.7 |
| *5<sup>th</sup> October 2020 – 14<sup>th</sup> February 2021 (Wave 2 - wild type)* |  |  |  |  |  |  |
| 1-4y | 17 | 22 | -5 | -22.7 | -38.8, 51.9 | -13.8 |
| 5-14y | 20 | 30 | -10 | -33.3 | -47.3, 12.3 | -10.9 |
| 15-44y | 79 | 84 | -5 | -6.0 | -14.5, 29.0 | -4.0 |
| 45-64y | 88 | 84 | 4 | 4.8 | -5.9, 42.8 | 12.4 |
| ≥65y | 200 | 180 | 20 | 11.1 | 7.0, 39.8 | 156.9 |
| All ages<sup>*</sup> | 404 | 400 | 4 | 1.0 | 0.0, 18.4 | 1.3 |
| *15<sup>th</sup> February 2021 – 4<sup>th</sup> June 2021 (Wave 3 – Beta-Alpha)* |  |  |  |  |  |  |
| 1-4y | 20 | 20 | 0 | 0.0 | -31.8, 92.1 | 0.0 |
| 5-14y | 21 | 21 | 0 | 0.0 | -27.0, 67.1 | 0.0 |
| 15-44y | 61 | 72 | -11 | -15.3 | -31.1, 6.6 | -8.8 |
| 45-64y | 75 | 78 | -3 | -3.8 | -19.6, 20.3 | -9.3 |
| ≥65y | 179 | 161 | 18 | 11.2 | -2.0, 29.7 | 141.2 |
| All ages<sup>*</sup> | 356 | 352 | 4 | 1.1 | -8.0, 12.2 | 1.3 |
| *5<sup>th</sup> June 2021 – 11<sup>th</sup> December 2021 (Wave 4 - Delta)* |  |  |  |  |  |  |
| 1-4y | 20 | 32 | -12 | -37.5 | -55.0, -10.5 | -33.2 |
| 5-14y | 19 | 41 | -22 | -53.7 | -65.7, -33.6 | -23.9 |
| 15-44y | 102 | 117 | -15 | -12.8 | -26.7, 4.1 | -12.0 |
| 45-64y | 156 | 120 | 36 | 30.0 | 9.0, 51.3 | 111.7 |
| ≥65y | 414 | 299 | 115 | 38.5 | 22.8, 57.1 | 902.1 |
| All ages<sup>*</sup> | 711 | 609 | 102 | 16.7 | 9.5, 25.6 | 34.3 |
| *12<sup>th</sup> December 2021 – 16<sup>th</sup> April 2022 (Wave 5 – Omicron BA1)<sup>†</sup>* |  |  |  |  |  |  |
| 1-4y | 19 | 21 | -2 | -9.5 | -36.8, 41.2 | -5.5 |
| 5-14y | 15 | 27 | -12 | -44.4 | -59.4, -14.9 | -13.1 |
| 15-44y | 71 | 81 | -10 | -12.3 | -26.9, 13.3 | -8.0 |
| 45-64y | 95 | 85 | 10 | 11.8 | -7.6, 34.1 | 31.0 |
| ≥65y | 207 | 177 | 30 | 16.9 | 2.1, 31.7 | 235.3 |
| All ages<sup>*</sup> | 407 | 391 | 16 | 4.1 | -5.9, 14.2 | 5.4 |

\*All ages excluding infants <1 year old
†After adjusting for under-ascertainment of deaths between 15th January – 5th May 2022.

On aggregate, across the first five waves of the pandemic, 1^st^ April 2020-16^th^ April 2022, we predicted 2350 deaths in adults and children (older than 1 year) based on 10 years of baseline data, but in fact we observed 2447 deaths, giving an excess mortality of 4.1% (95% PI -0.2%, 7.9%) and an excess mortality risk of 32.6/100,000 (Table 1b). Excess mortality only deviated significantly above zero among adults aged ≥65 years (14.3% 95% PI 7.4%, 21.6%); there was significantly negative excess mortality among children aged 1-4 years (−17.7%, 95% PI -30.5%, -2.5%) and 5-14 years (−22.4%, 95% PI -34.3%, -9.2%), and adults aged 15-44 years (−7.6%, 95% PI -14.9%, -0.2%) (Table 1b). Summary excess mortality in all children aged 1-14 years was -20.3% (95% PI -29.8%, -8.1%). In sex-specific analyses of the 5-wave period, overall excess mortality was similar for males (4.6%) and females (3.1%, Table S7a, Table S7b). However, among females, there was a significant excess mortality among those aged 45-64 years of 16.2% (95% PI 4.2%, 41.7%) and a significant negative excess among those aged 15-44 years of -17.0% (95% PI - 25.8%, -7.2%); the equivalent figures for males were -0.4% and 1.3%, respectively.

**Table 1b.** Accumulated excess deaths from 1^st^ Jan 2020 to 31 Dec 2021 in Kilifi HDSS residents aged >1 year.

| Age group | Deaths |  | Excess mortality |  |  |  |
| --- | --- | --- | --- | --- | --- | --- |
|  | observed | expected | N | % | 95% PI | Risk/100,000 |
| *1<sup>st</sup> April 2020 – 16<sup>th</sup> April 2022 (Waves 1-5)* |  |  |  |  |  |  |
| 1-4y | 107 | 130 | -23 | -17.7 | -30.5, -2.5 | -63.7 |
| 5-14y | 121 | 156 | -35 | -22.4 | -34.3, -9.2 | -38.1 |
| 15-44y | 437 | 473 | -36 | -7.6 | -14.9, -0.2 | -28.9 |
| 45-64y | 525 | 491 | 34 | 6.9 | -1.6, 16.9 | 105.5 |
| ≥65y | 1257 | 1100 | 157 | 14.3 | 7.4, 21.6 | 1231.6 |
| All ages* | 2447 | 2350 | 97 | 4.1 | -0.2, 7.9 | 32.6 |
| *1<sup>st</sup> January 2020 – 31<sup>st</sup> December 2021* |  |  |  |  |  |  |
| 1-4y | 107 | 128 | -21 | -16.4 | -31.6, -2.3 | -58.1 |
| 5-14y | 130 | 153 | -23 | -15.0 | -28.4, 0.1 | -25.0 |
| 15-44y | 442 | 464 | -22 | -4.7 | -12.1, 3.6 | -17.7 |
| 45-64y | 519 | 481 | 38 | 7.9 | 1.0, 17.0 | 117.9 |
| ≥65y | 1243 | 1066 | 177 | 16.6 | 9.7, 23.5 | 1388.5 |
| All ages* | 2441 | 2292 | 149 | 6.5 | 2.5, 9.6 | 50.1 |

\*All ages excluding infants <1 year old.

For comparison with modelled estimates of mortality we also calculated the excess mortality in Kilifi HDSS for the entire two-year period 1^st^ January 2020 – 31^st^ December 2022. Observed and expected deaths for all residents aged ≥1 year were 2441 and 2292 (Excess mortality 6.5%, 95% PI 2.5%, 9.6%); the excess mortality risk in this population was 50.1/100,000 persons during these two years (Table 1b). After standardizing the age-specific results from Kilifi HDSS to the age-structure of the Kenyan national population, the excess mortality risk was 47.5/100,000 persons during these two years.

There was no evidence that field interviewers were unable to reach appropriate respondents during the pandemic leading to under-reporting of deaths (Table S3). In survival analyses to 25^th^ October for HDSS cohorts that were resident on 23^rd^ March each year between 2010 and 2021, there was no evidence of decreased survival in 2020 (Table S4, Figure S2).

To explain the asynchrony between excess mortality peaks in Kilifi HDSS and the waves in the national case surveillance data, we analysed the subset of case surveillance data derived from Kilifi County. The surveillance data comprised PCR and rapid antigen tests for SARS-CoV-2, including negative results, collated across multiple Kenyan testing laboratories and national reporting mechanisms^30^. From 23^rd^ March 2020-5^th^ May 2022 Kilifi accounted for 79,310 (3.2%) of 2,501,682 PCR tests and 6,339 (1.1%) of 581,648 rapid antigen tests nationwide. Superimposing the national and Kilifi-specific epidemic curves (Figure S3) illustrates that wave 1 (wild-type) was absent in Kilifi, wave 3 (Beta-Alpha) arrived late in Kilifi and wave 4 (Delta) arrived early and was more pronounced.

### Cause of death (verbal autopsy)

Between 2015-2019, 71.7% (4647/6480) of deaths detected in Kilifi HDSS were investigated by verbal autopsy (Figure S4). The proportion of deaths investigated in 2020, 2021 and 2022 was 70.4% (914/1298), 63.5% (915/1440) and 47.2% (180/381), respectively; investigation of deaths occurring in 2022 is still ongoing. The proportion attributable to ARI was lower in 2020 in all age groups but returned to pre-COVID-19 levels in 2021 (Figure 3). The proportion attributable to RTA rose throughout the pandemic, particularly in those aged 15-44 years. The proportion attributable to stroke rose in 2021, particularly in those aged 45-64 years. Given approximately 1300 deaths per year, cause-specific mortality patterns are unstable over shorter intervals (Figure S5). However, the causes of death among adults aged ≥65 years in the three months prior to the pandemic did not differ from the causes in the remainder of 2020.

**Figure 3.**
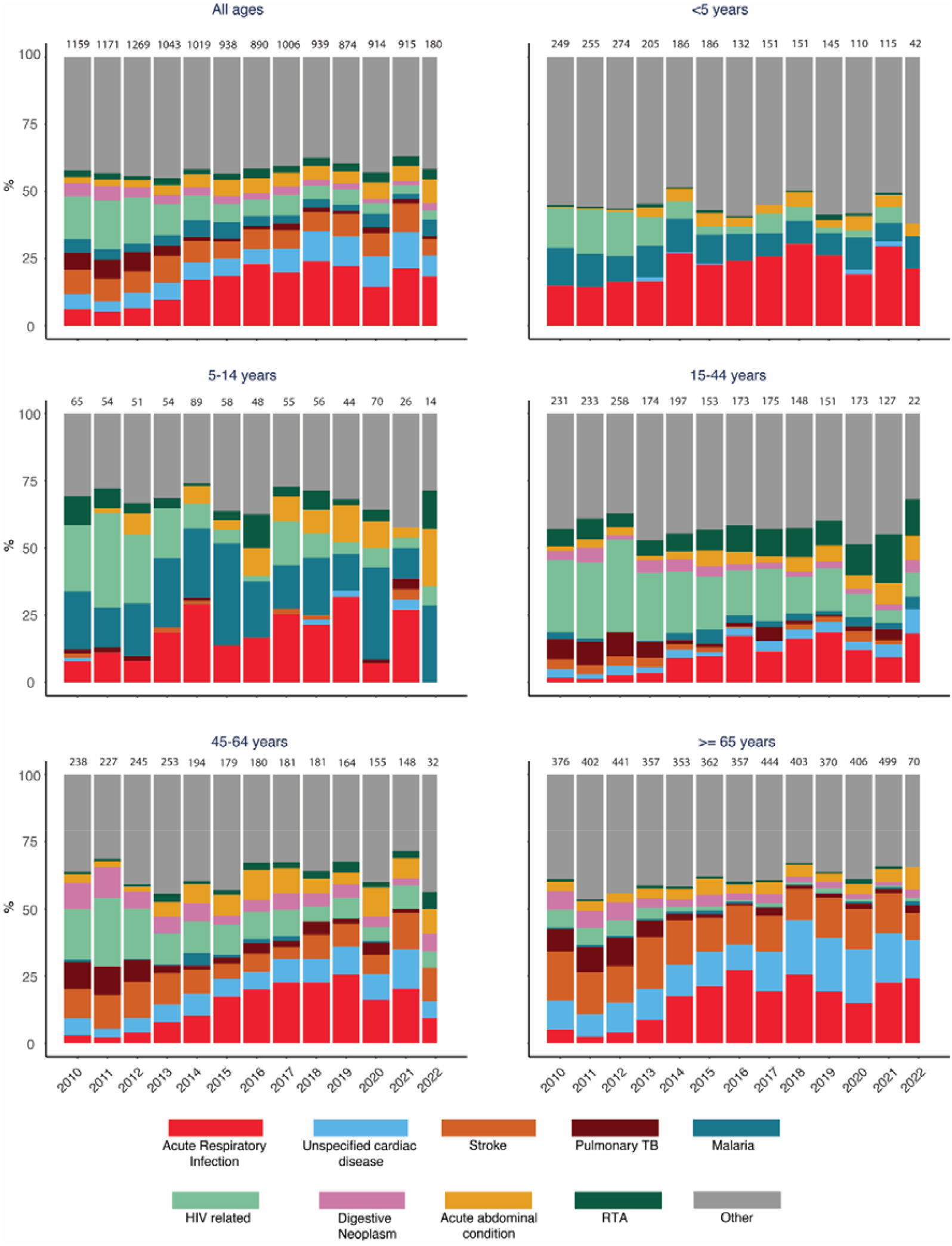
Annual cause-specific mortality fractions by Verbal Autopsy from 2010 to May 2022

Of 103 verbal autopsies where at least one COVID-19 specific answer was positive, 31 were defined as COVID-19 deaths using the algorithm, representing 1.8% of 1742 deaths investigated between 1^st^ April 2020 and 16^th^ April 2022. By physician review, 20 were probably related to COVID-19, 9 were possibly related and 74 were considered unrelated (Figure S6).

## DISCUSSION

The results of this longstanding mortality surveillance of a population of 306,000 in coastal Kenya indicate a mortality excess of 4.1% throughout the first five waves of the pandemic when compared to the 10 previous years. This aggregate figure conceals a higher mortality excess of 14.3% among those aged ≥65 years which is offset by a 20.3% reduction in mortality among children aged 1-14 years. Population excess mortality was only significantly positive during one of the five waves of SARS-CoV-2, the Delta wave; this is consistent with observations elsewhere that disease severity was greatest for the Delta variant^26,31,32^. The rise in seroprevalence of SARS-CoV-2 antibodies was much greater in 2021 than 2020 which may also explain the delayed impact on population mortality. Surprisingly, the second highest period of excess mortality was in the three months before the pandemic. Cause-specific analyses, using Verbal Autopsy, indicated a short-term reduction in the proportion of deaths caused by ARI in 2020; specific questions for COVID-19 identified only 31 deaths, implying poor sensitivity.

The strength of these findings is that they are direct empiric observations. Individuals in a large open cohort, who have been followed consistently for 20 years, were enumerated before the pandemic and re-enumerated repeatedly throughout the pandemic. Survival was verified by direct contact with the individual or by the report of a household member or close neighbour. Modelling was used only to generate a statistical fit to mortality data in the 10-year baseline period to provide a valid prediction of expected mortality during the pandemic period.

Nevertheless, the HDSS is susceptible to observation biases, particularly in March-October 2020 when fieldwork was suspended for 7 months. We may not have been able to ascertain all deaths among infants who were born during this suspension or among older persons who would normally have been expected to migrate into the area. We managed bias attributable to under-ascertainment of infant deaths by excluding infants from our analyses. We examined the potential of undetected immigrant deaths during March-October 2020 to obscure a COVID-19 attributable mortality excess among adults, by comparing the 7-month mortality experience of the resident cohorts at the suspension of fieldwork (23 March 2020) against the 7-month mortality experience of similar snapshot cohorts on this date in the previous 10 years; there was no evidence of an attenuation of survival during 2020 in these analyses.

Inferences may also be limited by instability in annual mortality rates in a population of 306,000, which is small by comparison with national datasets. We managed mortality variation in the baseline period by fitting our prediction model across 10 years of data. Reporting the mortality risk in the pandemic period after two years of observation further mitigated this risk of annual instability. Over smaller periods of time this instability is more problematic; the surprisingly high excess mortality observed during the first three months of 2020 may be an expression of this fluctuation.

The inaccessibility of COVID-19 testing in Africa and potential biases in the access to testing have led to controversy regarding the severity of COVID-19 in African populations^33^. An analysis of excess mortality cannot reliably estimate the infection fatality ratio for SARS-CoV-2 because it cannot distinguish the effects of the COVID-19 pandemic from those of the pandemic response. For example, the negative excess mortality in residents aged 1-44 years in Kilifi suggests there were net mortality benefits attributable to these restrictions; excess mortality could, therefore, substantially underestimate COVID-19-specific mortality in older adults, assuming deaths in young and old are similar and similarly ameliorated by pandemic restrictions.

In trying to discriminate these effects we found the VA data had limited value. There is a clear fall in the proportion of deaths due to ARI at all ages in 2020. This reverts to baseline levels in most age groups in 2021, whilst the pandemic waves continue, suggesting that pandemic restrictions in 2020 and adherence fatigue in 2021 are the more probable causes. Restrictions on movement during the pandemic can slow the transmission of existing respiratory pathogens^34^. Movement restrictions would also be expected to reduce the proportion of deaths attributable to road traffic accident but in Kilifi these increased, particularly among the age at highest risk (15-44 years); this paradoxical finding has also been observed in the USA^35^.

Among 1742 deaths examined between 1^st^ April 2020 and 16^th^ May 2022, the new WHO VA questions attributed only 31 (1.8%) to COVID-19 suggesting that the COVID-19 questions and algorithms lacked sensitivity. Deaths from COVID-19 may be mistaken for pneumonia deaths or they may be mediated through thrombotic pathology causing stroke or myocardial infarction^36^ and these pathways will not be captured by the COVID-19 VA questions. There is some evidence of an excess in the proportion of stroke deaths during 2021 among middle aged adults (45-64y, Figure 3).

The strongest peak in excess mortality, present at both 45-64 years and ≥65 years, occurred in July 2021. This timing may seem difficult to explain because it precedes the peak of the Delta wave in the national surveillance dataset by a few weeks. Sero-surveillance from the Kenya National Blood Transfusion service and modelling studies have illustrated marked regional heterogeneity in SARS-CoV-2 transmission patterns^30,37^ so we examined local COVID-19 testing data from Kilifi County. Although the data are relatively sparse, and susceptible to local ascertainment biases, they suggest that the rise in detected infections during wave 4 (Delta) in Kilifi precedes that in the national surveillance by approximately one month. The local data also help explain the absence of a mortality excess during the first wave (wild-type) as there were few infections detected in Kilifi during this wave.

The negative excess mortality estimate of 20.3% among all children aged 1-14 years is a large and surprising observation which may be attributable to social restrictions and school closures, limiting the spread of respiratory and gastrointestinal pathogens. Among young children, aged 1-4 years, there is a single peak of excess mortality coincident with the national Beta-Alpha peak but, for the rest of the pandemic, mortality in this age group was lower than predicted. Other studies of HDSS data from Western Kenya and Eastern Gambia have been unable to identify an excess of mortality in children aged<5 years^38^.

Whilst excess mortality cannot define the IFR for SARS-CoV-2, it can estimate the net mortality cost to society arising from the total experience of the pandemic. This is important for allocating resources between ongoing COVID-19 control measures and pre-existing health priorities. For Kenya, different global models have yielded widely varying results with profoundly different implications. The Global Burden of Disease models estimated excess mortality in Kenya over the first two years of the pandemic at 181.2/100,000^2^ whilst the World Health Organization^1^ model estimated an excess mortality of 11/100,000. For comparison, the modelled estimates for the USA were 179.3 and 140/10,000 respectively, and for the UK were 126.8 and 109/100,000, respectively. Our estimate of the net impact on mortality in the first two years of the pandemic is 47.5/100,000 after the Kilifi mortality data were standardised to the Kenya population structure. Of note, our estimate excludes the mortality experience of infants though, as noted above, there is little support for an excess of mortality among children in Africa. The COVID-19 pandemic might be more severe in urban than in rural settings^39^ and Kilifi HDSS has fewer urban residents (∼12%) than the national average (31%)^40^. It is possible, therefore, that data from Kilifi would underestimate the impact of the pandemic nationally. Nonetheless, these empiric data may be generalised to the 69% of the national population that live rural lives and they imply a pandemic mortality impact substantially lower than that observed in the United Kingdom or the USA.

This study provides observational data on the excess mortality risk associated with the pandemic in a population representative of much of rural tropical Africa. We are not aware of any previous observational studies in this region that document an increase in mortality risk. Whilst the mortality risk associated with the pandemic in coastal Kenya is substantially lower than that estimated in some global models it remains significant. Given that 25% of the adults in Kilifi HDSS had no measurable anti-SARS-CoV-2 antibodies when last sampled, a large proportion of this population remains susceptible to severe or fatal COVID-19 disease. Therefore, the evidence presented here, of a significant mortality risk among adults aged ≥65 years, supports a renewed focus on the delivery of COVID-19 vaccines to older persons in Kenya.

## Data Availability

Underlying individual data include geo-located residence and individual hospital records and hence would be high risk for identifiability. Intermediary data will be published on the Havard dataverse server under a CCBY 4.0 license.

## Declarations

### Ethics approval and consent to participate

Individual verbal consent to participate in a continuous health and demographic surveillance system was sought at the household level using a specific informed consent form. Written informed consent was obtained by interviewers from all VA respondents. This study was approved by the Ethical Review Committee of the Kenya Medical Research Institute (approval number: KEMRI/SERU/CGMR-C/007/3057).

### Authors’ contributions

Conceptualization: JAGS, TNW, PB. Data collection and preparation: AN, DW, GM, DA, CN. Formal analysis: MO, CB, JAGS Interpretation: MO, JAGS, CB, EWK, AOE, AIMO, EB, BT, JM, PB, AA, TNW. Writing, original draft preparation: MO, JAGS, CB. Writing, reviewing, and editing: all authors. Resources and funding acquisition: PB, JM, BT, JAGS, TNW. All authors critically reviewed the article and approved the final version for submission.

## Acknowledgements

We gratefully thank the residents of Kilifi who have participated in the surveillance activities of the Kilifi HDSS. We acknowledge the tremendous work of the verbal autopsy and census field staff, and data supervisors who collect and process this information, and the Community Liaison Group who run the community engagement programmes. This article is published with the permission of the Director of the Kenya Medical Research Institute.

