## Supplementary file for "Impact of COVID-19 on mortality in coastal Kenya: a longitudinal open cohort study"

Impact of the COVID-19 epidemic on mortality in Rural Coastal Kenya

**Authors and affiliations**

Appendix 1

Statistical models

**Equation S1: Negative binomial model for mortality**

$$y_{t}∽Negative Binomial \left( \mu_{t},\phi\right),$$

$$\log\mu_{t}=\beta_{0}+\beta_{1}t+\beta_{2}\cos\left( \frac{2\pi t}{12} \right)+\beta_{3}\sin\left( \frac{2\pi t}{12} \right)+\log\gamma_{t}.$$

$$y_{t}=no. of deaths per month$$

The model includes terms to account for log-linear trend and seasonality (sine and cosine terms) and an offset ($\log\gamma_{t})$to account for changes in person years of observation.

### **Table S1**. Additional VA questions for identification of possible COVID-19 deaths

| **ID** | **Question** | **Responses** |
| --- | --- | --- |
| id10482 | Was there any diagnosis by a health professional of COVID-19? | yes; no; don’t know; refused to answer |
| id10483 | Did he/she have a recent test by a health professional for COVID-19? | yes; no; don’t know; refused to answer |
| id10484 | What was the result? | positive; negative; unclear; don’t know; refused to answer |
| id10485 | Did he/she suffer from extreme fatigue? | yes; no; don’t know; refused to answer |
| id10486 | Did he/she experience a new loss, change or decreased sense of smell or taste? | yes; no; don’t know; refused to answer |
| id10487 | In the two weeks before death, did he/she live with, visit, or care for someone who had any COVID-19 symptoms, or a positive COVID-19 test? | yes; no; don’t know; refused to answer |

### **Table S2.** Timeliness of ascertainment of deaths by re-enumeration round

| **Re-enumeration round (n)** | **start date** | **finish date** | **all deaths detected by date of occurrence** | **deaths detected in round**  **n** | **% detected in round**  **n** | **deaths detected in round n+1** | **% detected in round n+1** | **deaths detected after round n+1** | **%. detected after round n+1** |
| --- | --- | --- | --- | --- | --- | --- | --- | --- | --- |
| 37 | 16/11/2015 | 02/05/2016 | 611 | 400 | 65.5 | 197 | 32.2 | 14 | 2.3 |
| 38 | 03/05/2016 | 23/08/2016 | 428 | 225 | 52.6 | 198 | 46.3 | 5 | 1.2 |
| 39 | 24/08/2016 | 09/01/2017 | 440 | 207 | 47.0 | 233 | 53.0 | 0 | 0.0 |
| 40 | 10/01/2017 | 13/07/2017 | 698 | 432 | 61.9 | 265 | 38.0 | 1 | 0.1 |
| 41 | 14/07/2017 | 09/01/2018 | 661 | 327 | 49.5 | 332 | 50.2 | 2 | 0.3 |
| 42 | 10/01/2018 | 30/04/2018 | 396 | 208 | 52.5 | 184 | 46.5 | 4 | 1.0 |
| 43 | 02/05/2018 | 03/09/2018 | 471 | 238 | 50.5 | 228 | 48.4 | 5 | 1.1 |
| 44 | 04/09/2018 | 03/01/2019 | 410 | 213 | 52.0 | 196 | 47.8 | 1 | 0.2 |
| 45 | 04/01/2019 | 23/04/2019 | 356 | 174 | 48.9 | 178 | 50.0 | 4 | 1.1 |
| 46 | 24/04/2019 | 18/08/2019 | 435 | 243 | 55.9 | 181 | 41.6 | 11 | 2.5 |
| 47 | 19/08/2019 | 02/01/2020 | 442 | 223 | 50.5 | 214 | 48.4 | 5 | 1.1 |
| 48 | 03/01/2020 | 11/01/2021 | 1305 | 556 | 42.6 | 734 | 56.2 | 15 | 1.1 |
| 49 | 12/01/2021 | 03/05/2021 | 375 | 199 | 53.1 | 172 | 45.9 | 4 | 1.1 |
| 50 | 04/05/2021 | 23/09/2021 | 601 | 313 | 52.1 | 288 | 47.9 |  |  |
| 51 | 24/09/2021 | 14/01/2022 | 247 | 247 |  |  |  |  |  |
| 37-47 | 16/11/2015 | 02/01/2020 | 5348 | 2890 | 54.0 | 2406 | 45.0 | 52 | 1.0 |

Baseline ascertainment was assessed for rounds 37-47 because, in subsequent rounds, the two-round lag period required to ascertain all deaths was interrupted by the pandemic (R48-49). This summary is based on events captured on or before 15^th^ August 2022.

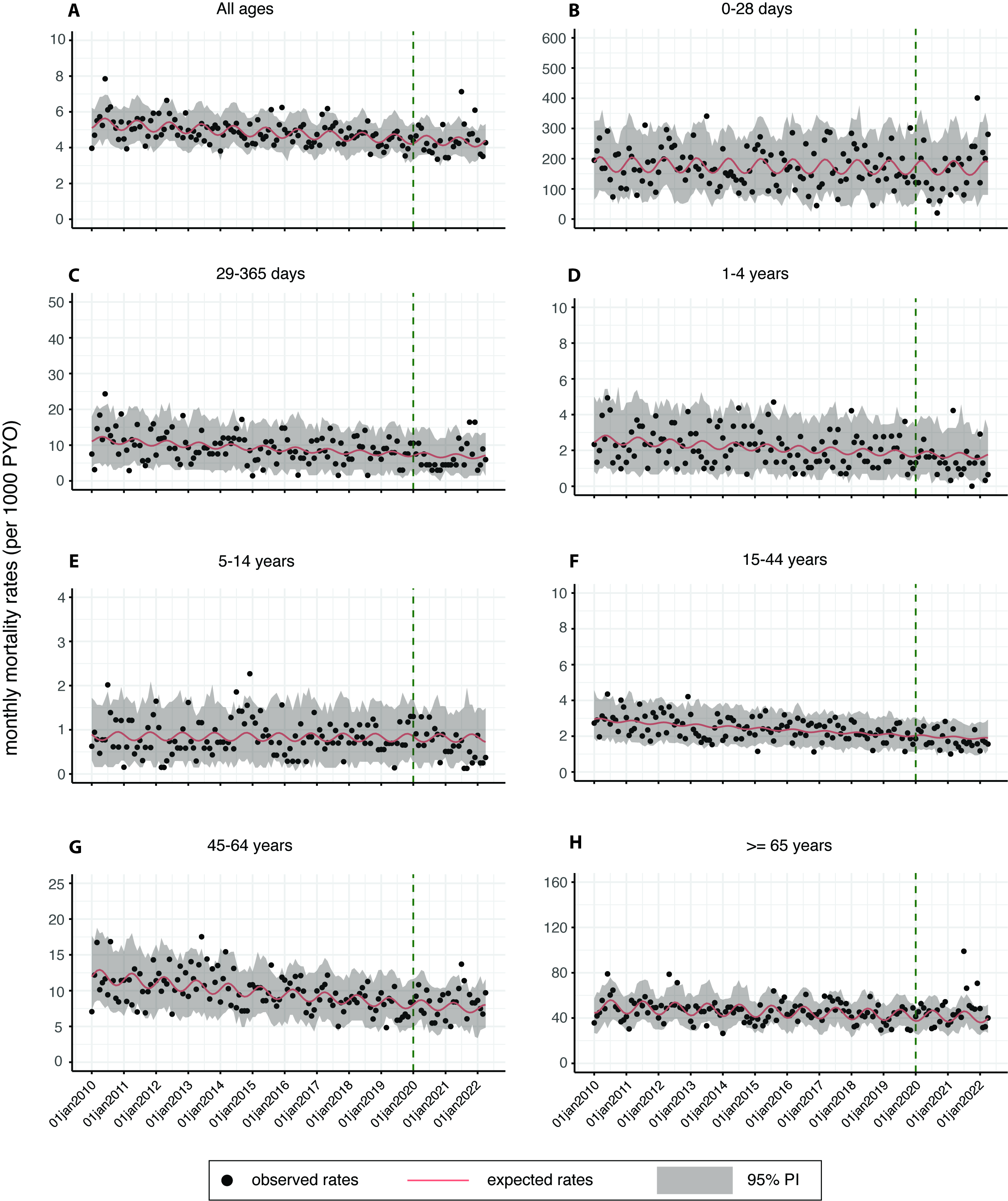

### **Figure S1.** Observed and expected monthly death rates from 01 January 2010 - 30 April 2022. Expected deaths rates were estimated from a negative binomial regression fitted using data up to 2019

Appendix 2 Data Quality Checks

Because there is a theoretical possibility of under-ascertainment of deaths during the period 23 March 2020 to 25 October 2020, when Kilifi HDSS field operations were suspended, we performed two data quality checks to investigate this possibility.

Firstly, we examined the source of information on deaths, current residents and out-migrants. If a fieldworker cannot find a household member during a re-enumeration round, the interview may be conducted with a respondent from a neighbouring household. One possibility to explain under-ascertainment of deaths during the pandemic period is that the fieldwork during this period relied more heavily on external respondents. We looked at the proportion of reports given by a respondent who was a resident of the same household.

Table S3 illustrates the variation in this proportion over the 13-year period of the mortality analysis. Deaths were reported by a member of the same household within a range of 71-83% in the period before fieldwork was suspended by Government restrictions on 23^rd^ March 2020. After fieldwork resumed on October 26^th^ there was no evidence of an increase in unreliable, external witnesses; in fact, the reporting of deaths was more likely to originate from household members than during the baseline period, probably because of reduced mobility and a greater likelihood of finding a respondent at home

### **Table S3.** Distribution of Kilifi HDSS respondents by time period of fieldwork.

|  | % of events reported by  members within the household | | |
| --- | --- | --- | --- |
| Year | deaths | outmigrations | enumerations |
| 2010 | 83 | 89 | 98 |
| 2011 | 80 | 86 | 97 |
| 2012 | 78 | 86 | 98 |
| 2013 | 81 | 84 | 98 |
| 2014 | 72 | 84 | 96 |
| 2015 | 72 | 84 | 96 |
| 2016 | 73 | 90 | 97 |
| 2017 | 80 | 87 | 98 |
| 2018 | 75 | 85 | 97 |
| 2019 | 71 | 86 | 96 |
| 2020 (Jan - Mar) | 78 | 87 | 97 |
| 2020 (Oct-Dec) | 79 | 81 | 95 |
| 2021 | 81 | 84 | 96 |
| 2022 (01 Jan - 05 May) | 72 | 84 | 96 |

Secondly, we explored the possibility that travel restrictions may have reduced a phenomenon known as the unhealthy immigrant; this is a scenario where a sick person returns to his or her rural home to die. This contributes to all-cause mortality in Kilifi HDSS but such sick individuals, about to die, may not have been able to travel to Kilifi during the pandemic period. If this effect was substantial, the unhealthy immigrant deaths unobserved during the pandemic period may have masked an excess of COVID-19 related deaths among stable residents of Kilifi HDSS. To explore this potential bias we calculated the 7-month risk of death in a cohort of residents selected on 23^rd^ March 2020, the day after the first case of COVID-19 was detected in Kilifi, and compared this cohort to similar cohorts selected on the same date in previous years. Table S4 shows the annual mortality risks. Figure S5 shows the survival of each cohort from 2010-2020. These analyses illustrate the mortality risk and survival function after excluding any immigrants during the risk period. The results do not suggest an excess of mortality risk or reduced survival in 2020/21 among stable residents of the Kilifi HDSS.

### **Table S4.** Age-adjusted 7-month risk of death in cohorts of residents selected on 23 March of each year from 2010-2021

| **Cohort year** | **All ages** | | **<65 years** | | **≥65 years** |  |
| --- | --- | --- | --- | --- | --- | --- |
|  | **Population** | **7-month mortality risk (%)** | **Population** | **7-month mortality risk (%)** | **Population** | **7-month mortality risk (%)** |
| 2010 | 248,095 | 0.44 | 240,016 | 0.27 | 8,079 | 4.79 |
| 2011 | 253,851 | 0.35 | 245,177 | 0.22 | 8,674 | 3.71 |
| 2012 | 258,706 | 0.36 | 249,780 | 0.22 | 8,926 | 3.98 |
| 2013 | 266,809 | 0.35 | 257,740 | 0.23 | 9,069 | 3.65 |
| 2014 | 271,235 | 0.34 | 261,718 | 0.22 | 9,517 | 3.42 |
| 2015 | 275,538 | 0.35 | 265,589 | 0.23 | 9,949 | 3.69 |
| 2016 | 278,585 | 0.32 | 267,661 | 0.20 | 10,924 | 3.42 |
| 2017 | 285,035 | 0.38 | 273,950 | 0.23 | 11,085 | 4.56 |
| 2018 | 289,863 | 0.31 | 278,163 | 0.20 | 11,700 | 3.33 |
| 2019 | 296,009 | 0.28 | 284,049 | 0.17 | 11,960 | 3.06 |
| 2020 | 300,671 | 0.30 | 288,008 | 0.19 | 12,663 | 3.20 |
| 2021 | 300,860 | 0.32 | 287,702 | 0.18 | 13,158 | 4.03 |

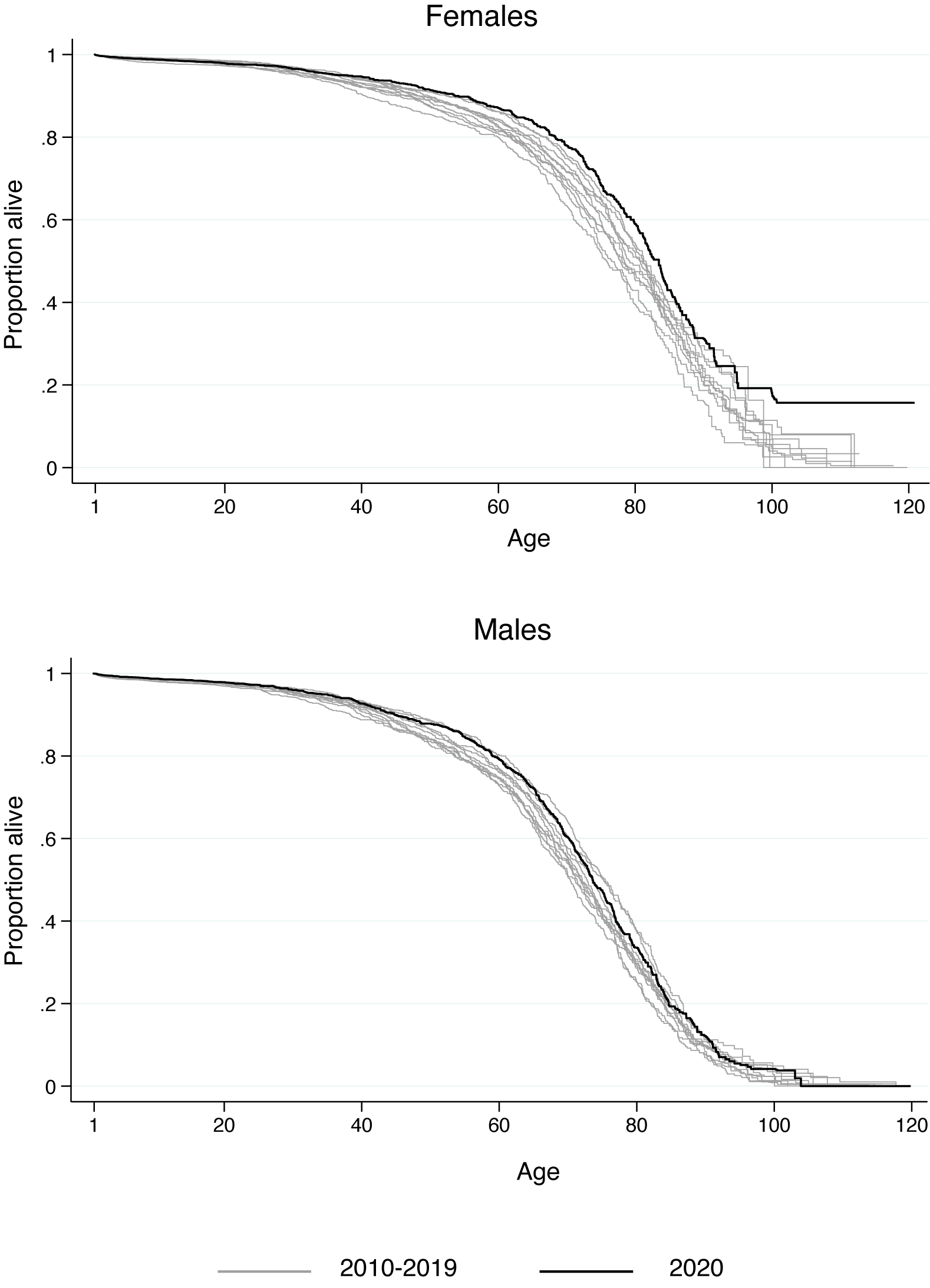

### **Figure S2**. Annual period survival curves for cohorts resident on the 23^rd^ March each year from 2010-2020 for males and females. Survival time is age, and the survival function starts at age 1 year, for consistency with other analyses. The light grey lines are individual survival curves of cohorts for the years 2010-2019 and the black solid lines are the survival curves for the 2020 cohort.

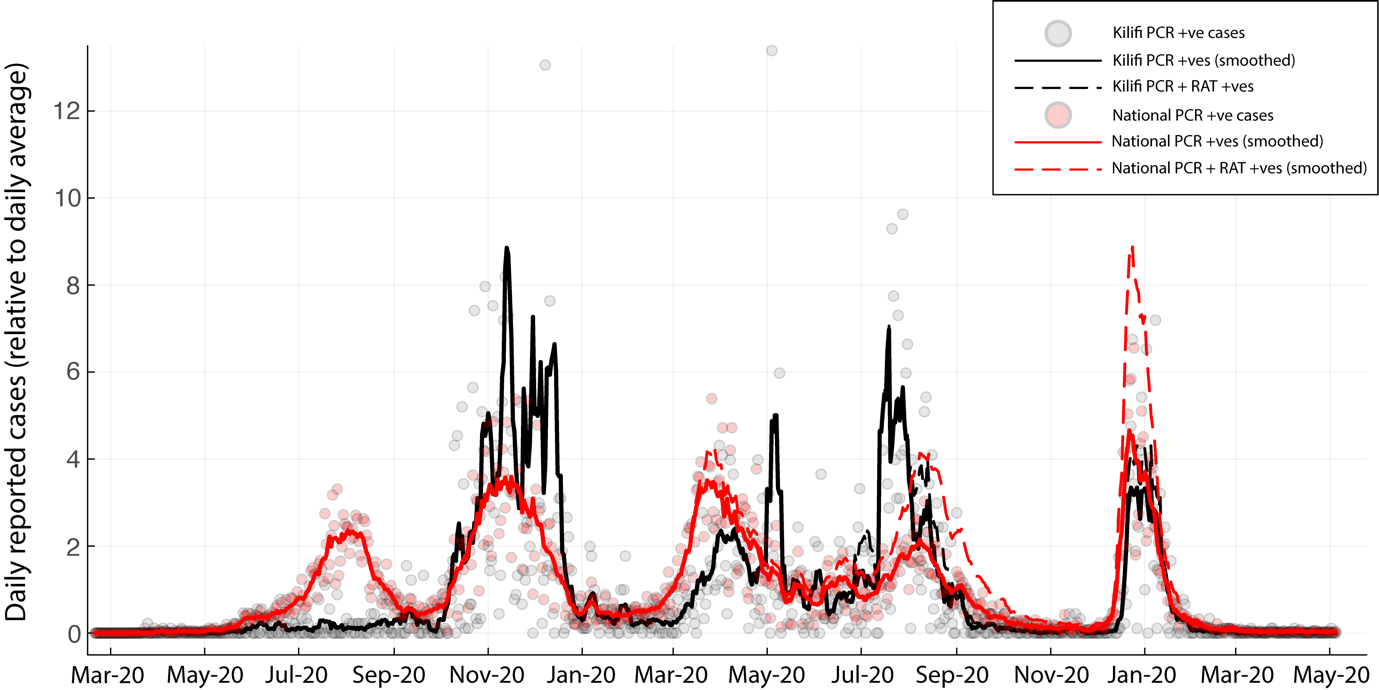

**Figure S3.** Ratio of COVID-19 cases reported daily to the mean daily number in National COVID-19 surveillance and the subset of data from Kilifi County alone. Initially, cases were defined by PCR testing alone. After DD/MM/2021 rapid antigen testing was introduced into the surveillance system.

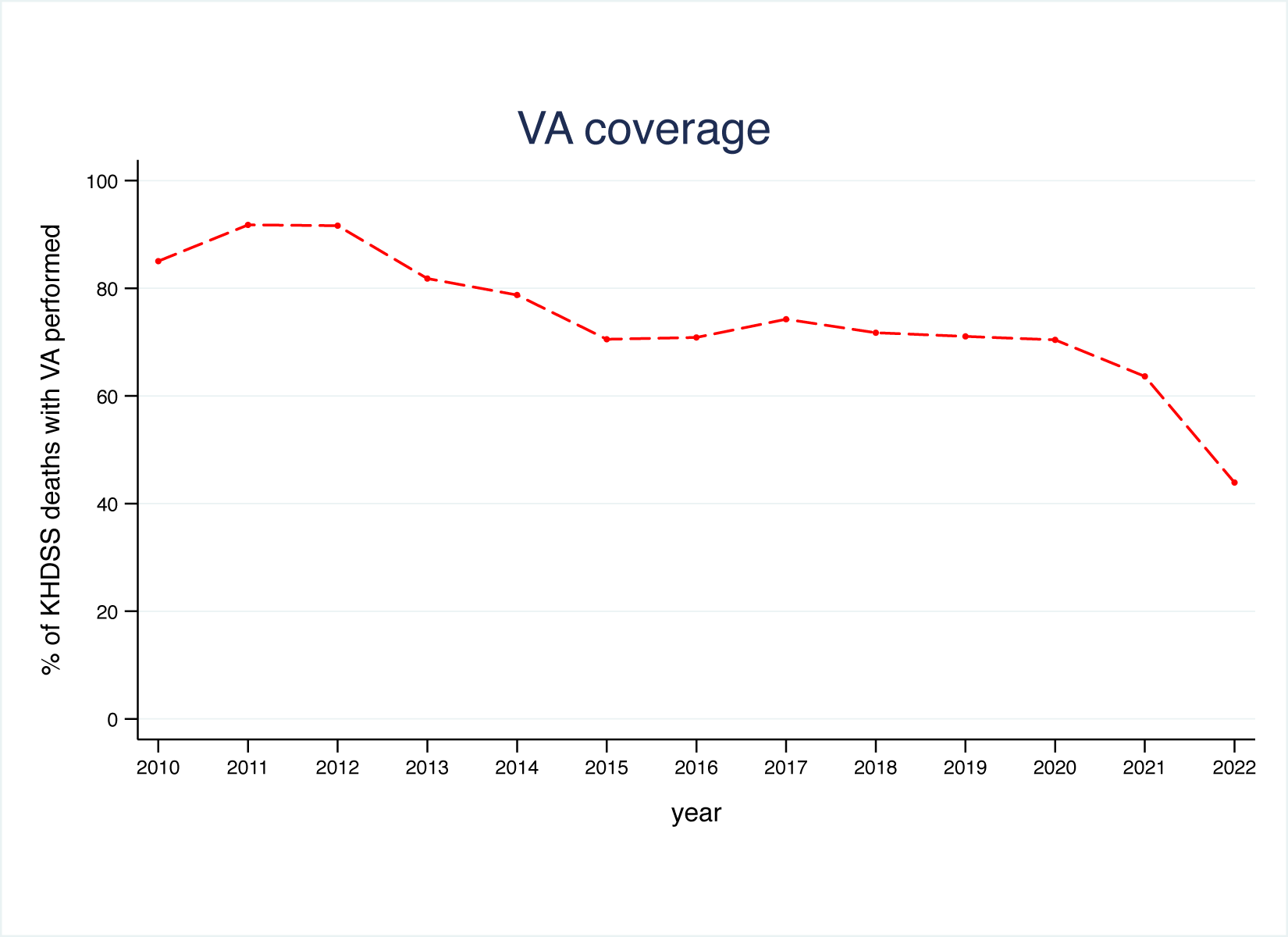

### **Figure S4** The proportion of all deaths recorded in Kilifi Health and Demographic Surveillance System that are investigated by verbal autopsy during the baseline period 2010-2019.Between 1st January 2020 and 31st December 2021, 1822 (66%) of 2736 deaths were investigated by verbal autopsy. Reasons for incomplete investigation include; inappropriate respondent, respondent not at home, postponed interview or refusal by the respondent. Despite longer intervals between death and ascertainment of death in 2020/21 finding an appropriate respondent was only marginally more difficult

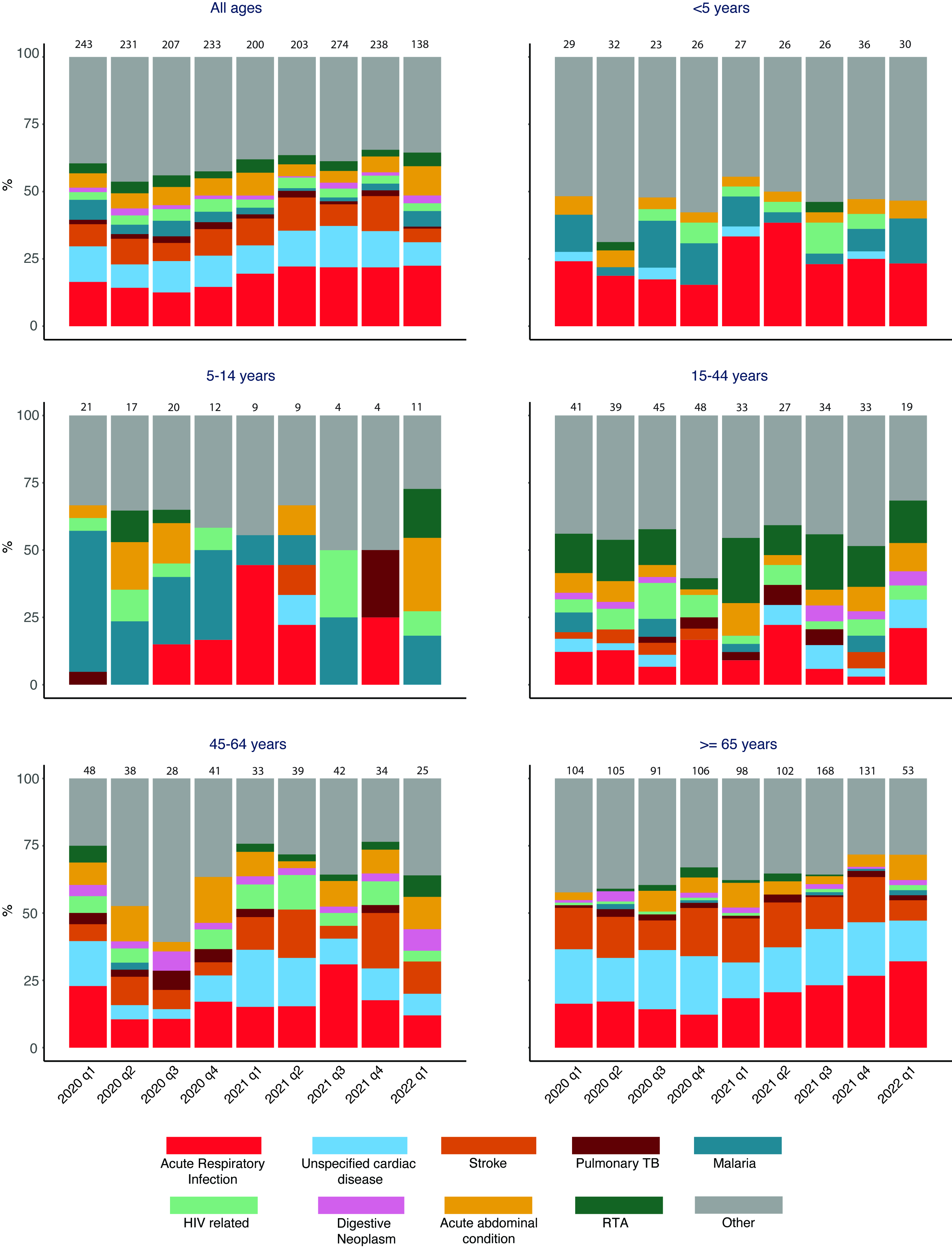

### **Figure S5** Quarterly cause-specific mortality fractions by Verbal Autopsy from January 2020 to March 2022. The excess mortality among adults aged ≥65 years in the first quarter of 2020 is not accompanied by a change in cause-specific patterns.

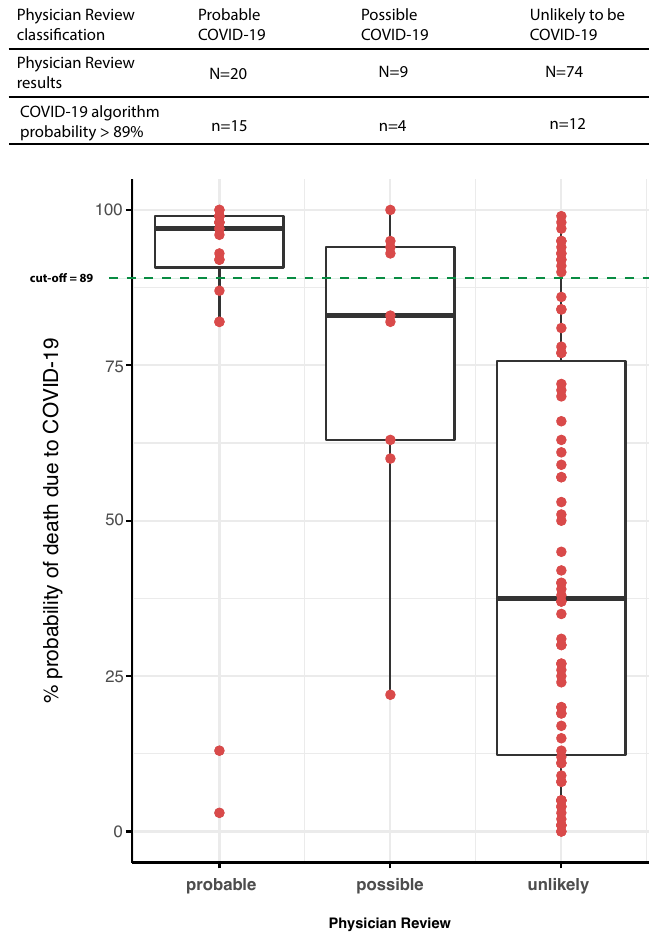

### **Figure S6.** Verbal Autopsy Assignment of COVID-19 as cause of death by CRMS algorithm and by physician review. Red dots are the probability values from the CRMS algorithm. They have been overlaid on their respective box plots which shows their distribution. The plots have been categorized according to the physician review categories. The green horizontal dotted line shows the probability cut-off value (89%) beyond which a death is classified as COVID-19 related by the CRMS algorithm.

### **Table S5.** Exact dates of re-enumeration rounds, COVID-19 waves and excess mortality analysis periods as illustrated in Figure 1.

| Event | Start date | End date |
| --- | --- | --- |
| **Re-enumeration rounds** |  |  |
| Round 48 | 03-01-2020 | 11-01-2021 |
| Round 49 | 12-01-2021 | 03-05-2021 |
| Round 50 | 04-05-2021 | 23-09-2021 |
| Round 51 | 24-09-2021 | 14-01-2022 |
| Round 52 | 15-01-2022 | 05-05-2022 |
| Round 53 | 06-05-2022 | 15-09-2022 |
| **Wave duration** |  |  |
| Wave 1 (wild-type) | 15-04-2020 | 19-09-2020 |
| Wave 2 (wild-type) | 20-09-2020 | 30-01-2021 |
| Wave 3 (Beta-Alpha) | 31-01-2021 | 20-05-2021 |
| Wave 4 (Delta) | 21-05-2021 | 26-11-2021 |
| Wave 5 (Omicron BA1) | 27-11-2021 | 02-04-2022 |
| **Excess mortality analysis window** |  |  |
| Analysis 1 (wave 1) | 01-04-2020 | 04-10-2020 |
| Analysis 2 (wave 2) | 05-10-2020 | 14-02-2021 |
| Analysis 3 (wave 3) | 15-02-2021 | 04-06-2021 |
| Analysis 4 (wave 4) | 05-06-2021 | 11-12-2021 |
| Analysis 5 (wave 5) | 12-12-2021 | 16-04-2022 |

### Excess mortality analysis windows start 2 weeks after the start date of respective waves, except for wave 1 where the excess mortality starts 2 weeks after the first case was identified. Dates reported as *dd-mm-yyyy*

**Table S6a**. Excess deaths from 1^st^ January 2020 to 16^th^ April 2022 in **female** Kilifi HDSS residents aged ≥1y, pre-pandemic and by wave analysis period.

| **Age** | **Deaths** | | **Excess mortality** | | | |
| --- | --- | --- | --- | --- | --- | --- |
| **group** | **observed** | **expected** | **N** | **%** | **95% PI** | **Risk/100,000** |
| *1^st^ January 2020 – 31^st^ March 2020 (pre-pandemic period)* | | | | | |  |
| 1-4y | 9 | 8 | 1 | 12.5 | -38.0 , 200.0 | 2.8 |
| 5-14y | 7 | 8 | -1 | -12.5 | -53.3 , 105.6 | -1.1 |
| 15-44y | 31 | 29 | 2 | 6.9 | -16.2 , 72.2 | 1.6 |
| 45-64y | 28 | 26 | 2 | 7.7 | -22.2 , 64.7 | 6.2 |
| ≥65y | 69 | 58 | 11 | 19 | -7.4 , 43.8 | 86.3 |
| All ages^*^ | 144 | 129 | 15 | 11.6 | -5.4, 27.1 | 5.0 |
| *1^st^ April 2020 – 4^th^ October 2020 (Wave 1 - wild type)* | | | | | |  |
| 1-4y | 12 | 15 | -3 | -20 | -47.2 , 38.7 | -8.3 |
| 5-14y | 25 | 17 | 8 | 47.1 | -4.6 , 149 | 8.7 |
| 15-44y | 60 | 60 | 0 | 0 | -17.3 , 29.4 | 0.0 |
| 45-64y | 53 | 55 | -2 | -3.6 | -22.6 , 27.6 | -6.2 |
| ≥65y | 120 | 137 | -17 | -12.4 | -22.8 , 0.3 | -133.4 |
| All ages^*^ | 270 | 284 | -14 | -4.9 | -13.9, 4.3 | -4.7 |
| *5^th^ October 2020 – 14^th^ February 2021 (Wave 2 - wild type)* | | | | | |  |
| 1-4y | 10 | 10 | 0 | 0 | -31.2 , 178.3 | 0.0 |
| 5-14y | 9 | 13 | -4 | -30.8 | -55.6 , 35.6 | -4.4 |
| 15-44y | 32 | 42 | -10 | -23.8 | -39.6 , 14.1 | -8.0 |
| 45-64y | 39 | 34 | 5 | 14.7 | -9.3 , 83.2 | 15.5 |
| ≥65y | 91 | 92 | -1 | -1.1 | -8.0 , 36.9 | -7.8 |
| All ages^*^ | 181 | 191 | -10 | -5.2 | -8.0, 13.9 | -3.4 |
| *15^th^ February 2021 – 4^th^ June 2021 (Wave 3 – Beta-Alpha)* | | | | | |  |
| 1-4y | 12 | 9 | 3 | 33.3 | -20.9 , 124.5 | 8.3 |
| 5-14y | 7 | 9 | -2 | -22.2 | -53.4 , 96.5 | -2.2 |
| 15-44y | 27 | 35 | -8 | -22.9 | -44.0 , 14.3 | -6.4 |
| 45-64y | 34 | 34 | 0 | 0 | -23.6 , 38.4 | 0.0 |
| ≥65y | 93 | 82 | 11 | 13.4 | -7.0 , 40.6 | 86.3 |
| All ages^*^ | 173 | 169 | 4 | 2.4 | -11.3, 13.6 | 1.3 |
| *5^th^ June 2021 – 11^th^ December 2021 (Wave 4 - Delta)* | | | | | |  |
| 1-4y | 7 | 14 | -7 | -50 | -67.5 , -16.0 | -19.4 |
| 5-14y | 9 | 19 | -10 | -52.6 | -69.2 , -13.3 | -10.9 |
| 15-44y | 44 | 59 | -15 | -25.4 | -40.6 , 6.7 | -12.0 |
| 45-64y | 77 | 51 | 26 | 51 | 22.1 , 103.3 | 80.7 |
| ≥65y | 214 | 153 | 61 | 39.9 | 23.0 , 61.6 | 478.5 |
| All ages^*^ | 351 | 296 | 55 | 18.6 | 8.4, 27.7 | 18.5 |
| *12^th^ December 2021 – 16^th^ April 2022 (Wave 5 – Omicron 1)* ^†^ | | | | | |  |
| 1-4y | 8 | 10 | -2 | -20 | -45.0 , 99.4 | -5.5 |
| 5-14y | 6 | 11 | -5 | -45.5 | -67.4 , 13.3 | -5.4 |
| 15-44y | 32 | 39 | -7 | -17.9 | -36.2 , 17.5 | -5.6 |
| 45-64y | 41 | 35 | 6 | 17.1 | -10.9 , 71.7 | 18.6 |
| ≥65y | 102 | 93 | 9 | 9.7 | -7.1 , 35.4 | 70.6 |
| All ages^*^ | 189 | 188 | 1 | 0.5 | -14.8, 13.2 | 0.3 |

*All ages excluding infants <1 year old

^†^After adjusting for under-ascertainment of deaths between 15th January – 5th May 2022.

**Table S6b**. Excess deaths from 1^st^ January 2020 to 16^th^ April 2022 in **male** Kilifi HDSS residents aged ≥1y, pre-pandemic and by wave analysis period.

| **Age** | **Deaths** | | **Excess mortality** | | | |
| --- | --- | --- | --- | --- | --- | --- |
| **group** | **observed** | **expected** | **N** | **%** | **95% PI** | **Risk/100,000** |
| *1^st^ January 2020 – 31^st^ March 2020 (pre-pandemic period)* | | | | | |  |
| 1-4y | 6 | 9 | -3 | -33.3 | -60.0 , 76.2 | -8.3 |
| 5-14y | 15 | 11 | 4 | 36.4 | -11.8 , 275.0 | 4.4 |
| 15-44y | 32 | 30 | 2 | 6.7 | -25.6 , 56.4 | 1.6 |
| 45-64y | 43 | 37 | 6 | 16.2 | -11.4 , 72.6 | 18.6 |
| ≥65y | 72 | 58 | 14 | 24.1 | 3.6 , 71.4 | 109.8 |
| All ages^*^ | 168 | 145 | 23 | 15.9 | 2.8, 32.4 | 7.7 |
| *1^st^ April 2020 – 4^th^ October 2020 (Wave 1 - wild type)* | | | | | |  |
| 1-4y | 19 | 20 | -1 | -5.0 | -29.8, 68.0 | -2.8 |
| 5-14y | 21 | 20 | 1 | 5.0 | -29.2, 67.8 | 1.1 |
| 15-44y | 64 | 60 | 4 | 6.7 | -17.5, 43.2 | 3.2 |
| 45-64y | 58 | 69 | -11 | -15.9 | -33.4, 17.5 | -34.1 |
| ≥65y | 137 | 144 | -7 | -4.9 | -18.3, 11.9 | -54.9 |
| All ages^*^ | 299 | 313 | -14 | -4.5 | -16.2, 4.6 | -4.7 |
| *5^th^ October 2020 – 14^th^ February 2021 (Wave 2 - wild type)* | | | | | |  |
| 1-4y | 7 | 12 | -5 | -41.7 | -59.2 , 7.3 | -13.8 |
| 5-14y | 11 | 17 | -6 | -35.3 | -55.7 , 15.4 | -6.5 |
| 15-44y | 47 | 42 | 5 | 11.9 | -6.1 , 69.4 | 4.0 |
| 45-64y | 49 | 50 | -1 | -2.0 | -16.2 , 46.7 | -3.1 |
| ≥65y | 109 | 88 | 21 | 23.9 | 14.3 , 76.9 | 164.7 |
| All ages^*^ | 223 | 209 | 14 | 6.7 | 1.3, 27.8 | 4.7 |
| *15^th^ February 2021 – 4^th^ June 2021 (Wave 3 – Beta-Alpha)* | | | | | |  |
| 1-4y | 8 | 11 | -3 | -27.3 | -54.2 , 64.7 | -8.3 |
| 5-14y | 14 | 12 | 2 | 16.7 | -26.6 , 134.6 | 2.2 |
| 15-44y | 34 | 37 | -3 | -8.1 | -26.8 , 48.7 | -2.4 |
| 45-64y | 41 | 44 | -3 | -6.8 | -30.1 , 24.0 | -9.3 |
| ≥65y | 86 | 79 | 7 | 8.9 | -10.1 , 33.3 | 54.9 |
| All ages^*^ | 183 | 183 | 0 | 0.0 | -14.3, 11.8 | 0.0 |
| *5^th^ June 2021 – 11^th^ December 2021 (Wave 4 - Delta)* | | | | | |  |
| 1-4y | 13 | 18 | -5 | -27.8 | -53.2 , 23.0 | -13.8 |
| 5-14y | 10 | 23 | -13 | -56.5 | -67.2 , -30.3 | -14.2 |
| 15-44y | 58 | 59 | -1 | -1.7 | -23.0 , 27.0 | -0.8 |
| 45-64y | 79 | 68 | 11 | 16.2 | -3.9 , 51.8 | 34.1 |
| ≥65y | 200 | 146 | 54 | 37.0 | 17.0 , 55.7 | 423.6 |
| All ages^*^ | 360 | 314 | 46 | 14.6 | 1.3, 23.7 | 15.5 |
| *12^th^ December 2021 – 16^th^ April 2022 (Wave 5 – Omicron 1)* ^†^ | | | | | |  |
| 1-4y | 11 | 11 | 0 | 0.0 | -38.2 , 74.4 | 0.0 |
| 5-14y | 9 | 15 | -6 | -40.0 | -62.0 , -9.4 | -6.5 |
| 15-44y | 39 | 42 | -3 | -7.1 | -28.4 , 39.9 | -2.4 |
| 45-64y | 54 | 51 | 3 | 5.9 | -13.4 , 38.8 | 9.3 |
| ≥65y | 105 | 84 | 21 | 25.0 | 6.6 , 54.0 | 164.7 |
| All ages^*^ | 218 | 203 | 15 | 7.4 | -2.8, 19.1 | 5.0 |

*All ages excluding infants <1 year old

^†^After adjusting for under-ascertainment of deaths between 15th January – 5th May 2022.

**Table S7a**. Excess deaths in waves 1-5, and from 1^st^ Jan 2020 to 31 Dec 2021 in Kilifi HDSS residents aged >1 year in females.

| **Age** | **Deaths** | | **Excess mortality** | | | |
| --- | --- | --- | --- | --- | --- | --- |
| **group** | **observed** | **expected** | **N** | **%** | **95% PI** | **Risk/100,000** |
| *1^st^ April 2020 – 16^th^ April 2022 (Waves 1-5)* | | | | | |  |
| 1-4y | 49 | 58 | -9 | -15.5 | -31.0 , 3.2 | -24.9 |
| 5-14y | 56 | 69 | -13 | -18.8 | -35.0 , 5.0 | -14.2 |
| 15-44y | 195 | 235 | -40 | -17.0 | -25.8 , -7.2 | -32.1 |
| 45-64y | 244 | 210 | 34 | 16.2 | 4.2 , 41.7 | 105.5 |
| ≥65y | 620 | 557 | 63 | 11.3 | 2.0 , 21.8 | 494.2 |
| All ages^*^ | 1164 | 1129 | 35 | 3.1 | -4.5, 5.6 | 11.8 |
| *1^st^ January 2020 – 31^st^ December 2021* | | | | | |  |
| 1-4y | 50 | 57 | -7 | -12.3 | -31.1 , 10.0 | -19.4 |
| 5-14y | 58 | 67 | -9 | -13.4 | -31.4 , 7.5 | -9.8 |
| 15-44y | 197 | 231 | -34 | -14.7 | -24.5 , -2.5 | -27.3 |
| 45-64y | 241 | 206 | 35 | 17.0 | 4.5 , 37.8 | 108.6 |
| ≥65y | 606 | 537 | 69 | 12.8 | 4.3 , 24.1 | 541.3 |
| All ages^*^ | 1152 | 1098 | 54 | 4.9 | -0.4, 10.0 | 18.2 |

*All ages excluding infants <1 year old

**Table S7b.** Excess deaths in waves 1-5, and from 1^st^ Jan 2020 to 31 Dec 2021 in Kilifi HDSS residents aged >1 year in males.

| **Age** | **Deaths** | | **Excess mortality** | | | |
| --- | --- | --- | --- | --- | --- | --- |
| **group** | **observed** | **expected** | **N** | **%** | **95% PI** | **Risk/100,000** |
| *1^st^ April 2020 – 16^th^ April 2022 (Waves 1-5)* | | | | | |  |
| 1-4y | 58 | 72 | -14 | -19.4 | -34.1 , -0.9 | -38.8 |
| 5-14y | 65 | 88 | -23 | -26.1 | -39.0 , -7.5 | -25.0 |
| 15-44y | 242 | 239 | 3 | 1.3 | -9.8 , 13.9 | 2.4 |
| 45-64y | 281 | 282 | -1 | -0.4 | -9.5 , 14.9 | -3.1 |
| ≥65y | 637 | 545 | 92 | 16.9 | 9.7 , 25.8 | 721.7 |
| All ages^*^ | 1283 | 1226 | 57 | 4.6 | -0.6, 11.0 | 19.2 |
| *1^st^ January 2020 – 31^st^ December 2021* | | | | | |  |
| 1-4y | 57 | 71 | -14 | -19.7 | -32.9 , -0.8 | -38.8 |
| 5-14y | 72 | 86 | -14 | -16.3 | -28.0 , 12.5 | -15.2 |
| 15-44y | 245 | 234 | 11 | 4.7 | -6.1 , 19.3 | 8.8 |
| 45-64y | 278 | 276 | 2 | 0.7 | -8.7 , 17.3 | 6.2 |
| ≥65y | 637 | 532 | 105 | 19.7 | 10.4 , 29.9 | 823.7 |
| All ages^*^ | 1289 | 1199 | 90 | 7.5 | 1.0, 14.8 | 30.3 |

*All ages excluding infants <1 year old
